# Quantifying the contribution of Oral Cholera Vaccine to global health through tech-transfers and WHO stockpile deployment since its inception: a modeling study

**DOI:** 10.64898/2026.02.02.26345439

**Authors:** Jung-Seok Lee, Wongyeong Choi, Florian Marks, John D. Clemens, Jerome H. Kim, Julia Lynch

## Abstract

**Introduction:** Cholera has been a major public health concern in many parts of the world. The development of the low-cost Oral Cholera Vaccine (OCV) enabled creation of the OCV stockpile by the World Health Organization and Gavi, the Vaccine Alliance which have provided vaccine to cholera-affected countries since 2013. The current study aims to measure the impact of the OCV stockpile deployment on global health.

**Methods:** Detailed OCV stockpile data were obtained and used to determine the size and timing of vaccination using a transmission model across all recipient countries over time. The model first estimated the impact of vaccination based on actual shipments. Considering several challenges including time lags until vaccination and the shortage of OCVs observed during the OCV stockpile implementations, multiple scenarios, so called ‘what if scenarios’ were further investigated to provide guidance for the future decision-making processes on the OCV stockpile use.

**Results:** With the actual OCV shipment scenario, vaccination prevented 8.1 million cases and 146,013 deaths which resulted in 6.7 million DALYs averted. If there were neither time lags nor the supply shortages, vaccination would avert 17.4 million cases, 321,730 deaths, and 15.1 million DALYs. The economic burden of cholera reduced by vaccination was estimated to be US$1.05 billion with the actual OCV shipment and US$2.33 billion without having any constraints.

**Conclusion:** Our models indicate that deployment of OCV from the stockpile may have had a significant positive impact on cholera-affected countries since its inception. Overcoming existing challenges resulting in delays in campaign implementation and shortage of vaccine will help maximize the impact of vaccination.

**KEY MESSAGES:** *What is already known on this topic:* Cholera outbreaks are currently ongoing in multiple countries, with the African and Eastern Mediterranean regions experiencing the most severe impacts. The Oral Cholera Vaccine (OCV) stockpile was established by World Health Organization (WHO) and Gavi since 2013 and has been widely used to control cholera outbreaks globally. Existing modeling studies assessed the potential impact of OCV and reported that OCV campaigns can contribute to effective cholera control in the near term. However, while the global OCV stockpile has been available since 2013 supporting more than 30 cholera-endemic countries, the studies mostly focused on a specific time period in selected geographical locations, missing the holistic impact of the global OCV stockpile. We further reviewed existing daily OCV stockpile shipment records thoroughly and identified that the program faced several challenges in meeting urgent demands due to administrative delays and OCV supply shortages.

*What this study adds:* We evaluated the actual impact of deployed doses on cholera morbidity, mortality, and economic burden since the inception of the OCV stockpile in 2013. Our model further investigated what the impact of OCV would have been with no delays in implementation and no supply shortage by incorporating the detailed daily shipment data which included the number of OCV doses and dates for request, decision (approval), and delivery. The current analyses show that from 2013 to 2025, both reactive and preventive vaccination campaigns implemented through the global OCV stockpile averted more than 6.7 million DALYs, 146,013 deaths, and US$1.05 billion of the economic burden in 37 recipient countries. Our model demonstrates that vaccination impact would be further enhanced if there were no delays in the implementation of campaigns and if there were no shortage of OCV supplies.

*How this study might affect research, practice or policy:* To our knowledge, no previous study has assessed the holistic contribution of the global OCV stockpile and investigated the optimal outcomes which may have been partially lost due to programmatic and logistical challenges. Since its inception, the impact of the OCV stockpile on global health has been substantial, and the additional benefits are expected if cholera vaccines can be deployed on time and if there is no supply constraint. In particular, the benefits induced by improving timing of vaccination does not necessarily come with additional costs associated with vaccine supplies which are known to be the major cost driver. In addition, because all stockpile recipient countries are currently required to implement a single-dose OCV campaign due to the shortage of OCVs, the continuous support for a two-dose vaccination campaign would bring much larger benefits than the single-dose vaccination scheme given a single-dose vaccination with the OCV is known to be much less efficacious among young children. Considering ongoing cholera outbreaks across multiple regions and other cholera vaccine candidates in development, our findings can be used not only to help provide guidance for the decision-making process on the future OCV stockpile deployment, but also to understand the optimal usage and potential impacts of multiple cholera vaccines which can be administered by different strategies in the future.

## INTRODUCTION

Cholera is an acute diarrheal infection caused by ingestion of food or water contaminated with the bacterium *Vibrio cholerae*. While improvements in water, sanitation, and hygiene help mitigate disease transmission and prevent cholera infections, such efforts are considered to be long-term measures and challenging in the context of low- and middle-income country settings where resources are scarce. Several cholera vaccines already exist and have proved useful in effectively controlling the disease through both reactive and preemptive campaigns.

Dukoral, a two-dose inactivated vaccine, has been on market since 1991, but this vaccine has been mainly used in developed countries as a traveler’s vaccine due to its relatively high cost. In 1997, ORC-Vax, bivalent Oral Cholera Vaccine (OCV), was locally licensed in Vietnam.^1^ However, in order to meet the WHO requirements and be used at the global level, a team at the International Vaccine Institute (IVI) reformulated the vaccine in collaboration with VaBiotech in Vietnam, transferred the technology back to VaBiotech^1, 2^ and to Shantha Biotechnics in India. This vaccine, Shanchol, was prequalified by WHO in 2011 and became the world’s first low-cost OCV. Subsequently, workers at the IVI also transferred the same OCV technology to EuBiologics in Korea to help ensure a reliable supply for the global market. The WHO has coordinated a Gavi-supported OCV stockpile since 2013 that has provided Shanchol and Euvichol / Euvichol-Plus upon request by cholera-affected countries and availability for reactive and preventive campaigns. All three vaccines are killed whole cell vaccines and have the same strains and compositions (targeting O1 and O139 serogroups). While Shanchol is no longer available in the market, a simplified version of OCV, Euvichol-S, targeting only the O1 serogroup was recently prequalified by the WHO in April 2024.^3, 4^ The vaccine is indicated for persons aged 1 year or older with the recommended dosing schedule of two doses at least 14 days apart. It is known that two-doses given to persons at least five years of age provide substantial protection for at least 5 years^5^, but protection of children younger than 5 years is lower and lasts four years.^6, 7^ Current policies recommend a single dose vaccination in an outbreak setting where delivery of two doses is not possible, however the duration of protection may be reduced and no protection has been observed from a single dose in children under 5 years.^8^

The WHO OCV stockpile was initially designed for rapid outbreak responses or humanitarian crises, but with the availability of additional doses, it has been also used for preventive campaigns in communities at high risk (prioritized areas for cholera control). In other words, OCV is provided through geographically targeted campaigns, not through routine immunization. The emergency use of OCV aims to allow rapid access to vaccines to stop the spread of an outbreak. Though the emergency application procedures through the WHO International Coordinating Group (ICG) secretariat are designed to facilitate rapid response, a number of factors can delay implementation of campaigns and vaccination may occur after the peak of an outbreak which can result in minimal health impact. On the other hand, the preventive use of OCV is intended for targeted, well-planned campaigns focused on maximizing health impacts, but the application process takes a longer time as compared to emergency purposes. It should be noted that since the upsurge in global cholera outbreaks and a shortage of the OCV in 2022, a policy of prioritizing vaccines to outbreaks was established, resulting in virtually all doses being used for reactive rather than preemptive vaccination. In addition, some countries have received a smaller number of OCV doses than requested. Furthermore, all recipient countries were asked to implement a single-dose vaccination campaign since October 2022, which in turn, has the potential to reduce the impact of the vaccination.

Since the first shipment from the OCV stockpile in 2013, more than 30 cholera-affected countries have received OCVs. The main aim of the current study was to model the actual impact of deployed doses on cholera morbidity, mortality, and economic burden. The study further investigated what the impact of OCV would have been if there had been no delays in the implementation of vaccination campaigns and if the campaigns had not been constrained by shortages of OCV doses in order to help provide guidance for the future decision-making process on the OCV stockpile use.

## METHODS

### OCV stockpile

To quantify the impact of the OCV stockpile, the number of OCV stockpile doses which had been shipped from its inception (2013) to 2025 were obtained from the Global Task Force on Cholera Control.^9^ The detailed daily shipment data including the number of OCV doses and dates for request, decision (approval), and delivery were manually extracted from the system by country and mechanism (preventive, reactive) over time.

### Model structure and calibration

An ordinary differential equation transmission model was constructed with multiple compartments including bacteria in water where two climate indicators (temperature, precipitation) were incorporated as shown in Supplementary Figure 1. In addition, two vaccination compartments were considered to tease out the vaccination impact between a single-dose and two-dose vaccinations (see the supplementary material for full details about the model structure).

Key parameters are summarized in Table 1 (see Supplementary Table 1 and Table 2 for further details). The model was first calibrated based on country-specific incidences which were derived from the case fatality rates^10^ and the number of deaths due to cholera^11^ for all 37 countries which had received the OCV stockpiles since 2013. While a large proportion of cholera cases is harbored in high-risk areas, it is also possible that cholera cases can be identified in medium- to low-risk areas. The current study presumed that the OCV stockpile was prioritized to high-risk areas, which resulted in effective prevention against cholera in the areas of vaccination but still leaving medium- to low-risk areas uncovered within a country. In order to estimate the proportion of high-risk areas in each country, a composite index called the typhoid risk-factor (TRF) index was adopted^12^, considering that typhoid and cholera diseases share some of the common risk factors such as drinking water sources, hygiene facilities, and population density.

**Table 1.**
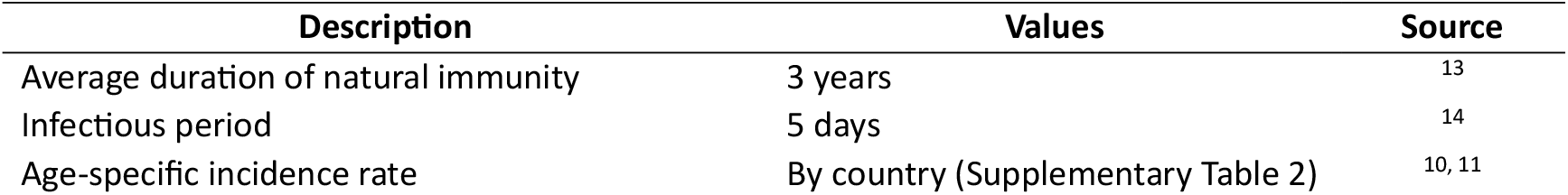
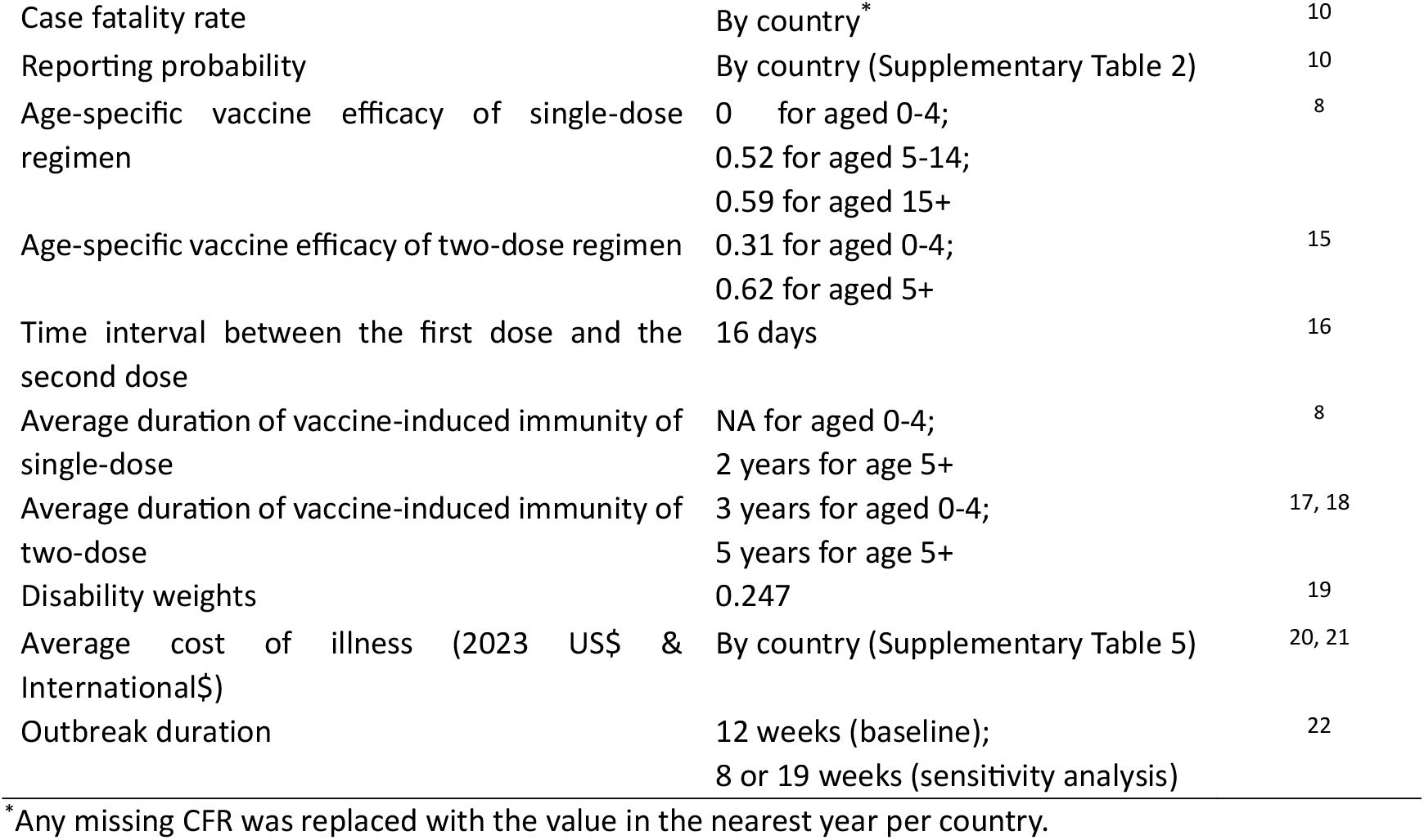
Key parameters and data.

| Description | Values | Source |
| --- | --- | --- |
| Average duration of natural immunity | 3 years | 13 |
| Infectious period | 5 days | 14 |
| Age-specific incidence rate | By country (Supplementary Table 2) | 10, 11 |
| Case fatality rate | By country* | 10 |
| Reporting probability | By country (Supplementary Table 2) | 10 |
| Age-specific vaccine efficacy of single-dose regimen | 0 for aged 0-4;<br>0.52 for aged 5-14;<br>0.59 for aged 15+ | 8 |
| Age-specific vaccine efficacy of two-dose regimen | 0.31 for aged 0-4;<br>0.62 for aged 5+ | 15 |
| Time interval between the first dose and the second dose | 16 days | 16 |
| Average duration of vaccine-induced immunity of single-dose | NA for aged 0-4;<br>2 years for age 5+ | 8 |
| Average duration of vaccine-induced immunity of two-dose | 3 years for aged 0-4;<br>5 years for age 5+ | 17, 18 |
| Disability weights | 0.247 | 19 |
| Average cost of illness (2023 US\$ & International\$) | By country (Supplementary Table 5) | 20, 21 |
| Outbreak duration | 12 weeks (baseline);<br>8 or 19 weeks (sensitivity analysis) | 22 |

\*Any missing CFR was replaced with the value in the nearest year per country.

Given a scarcity of the longitudinal data for cholera cases among the OCV stockpile recipient countries, it was challenging to understand the accurate size of various cholera outbreaks that each country had experienced over time since 2013. In the current study, the Cholera Platform and a previous study which reported weekly cholera cases longitudinally were used for the model to mimic the size of the observed outbreaks. The Regional Cholera Platform run by United Nations Office for the Coordination of Humanitarian Affairs provides weekly cholera cases from January 2012 to December 2017 for 18 countries in west and central Africa, in which ten countries were identified as the WHO OCV recipients.^23, 24^ In addition, a previous study provided cholera cases from October 2010 to January 2019 for Haiti.^25^ The observed outbreaks were plotted over time, and unusual peaks were categorized into three groups based on its relative outbreak size and used to calculate three multipliers corresponding to the three outbreak groups: outbreak 1 (large), outbreak 2 (medium), and outbreak 3 (small). When an outbreak was observed in the existing data^23-25^, one of the multipliers matching to the outbreak was applied to a country-specific reproduction number (R_0_) to approximate the size of the observed outbreak. The outbreak sizes were then verified again based on the number of the observed cholera cases.

In order to determine the size of outbreaks for all OCV recipient countries during the entire study period, the proportion of the reactive OCV doses requested out of the at-risk population was used. If the proportion of a single reactive OCV request relative to the risk population was (1) greater than the 75^th^ percentile, (2) between the 25^th^ percentiles and the 75^th^ percentiles, and (3) lower than the 25^th^ percentiles among all the requests, the size of an outbreak was assumed to be large (outbreak 1), medium (outbreak 2), and small (outbreak 3), respectively.

### Scenarios and input values

A previous study reported the median time lag from the initiation of an event to the time of actual vaccination for the global OCV use.^16^ As shown in the upper panel of Figure 1, it takes about 26 days from the time of an event to an OCV request, 5 days to get the request approved, 13 days to get OCVs delivered, and additional 15 days to have actual vaccination occur, resulting in a total of 59 days of a time lag in median for an outbreak situation. In addition to the existing evidence, considering that actual time lags between OCV requests and deliveries are variable by country and request, the current model was constructed using the country-specific observed request and delivery dates for each shipment to reflect the lags in a more realistic manner (the lower panel of Figure 1).

For vaccination impact estimations, four scenarios were considered and summarized in Supplementary Table 4. The first scenario is called ‘As what has happened’ which basically reflects the actual OCV shipment where there were three empirical constraints: (1) time lags from the time of an outbreak event to the time of vaccination (for a preventive vaccination, from the time of a request to vaccination), (2) some countries receiving less number of OCV doses than what they requested due to OCV supply shortages, (3) requiring all recipient countries to implement only a single-dose vaccination campaign since October 2022, again due to the OCV supply limitation. The second scenario then relaxes the first constraint, thus considers what if there were no time lags between the time of an event and the time of vaccination, holding the other constraints same. To estimate the impact of vaccination without the supply shortage independent of the other constraints, the third scenario loosens the constraints (2) and (3) – what if there were enough OCV supplies meeting all the requests, holding the first constraint (time lags) constant. Finally, the fourth scenario eases all three constraints and investigates what the vaccination impact would have been if there had been no time lags with enough OCV supplies as requested.

**Figure 1.**
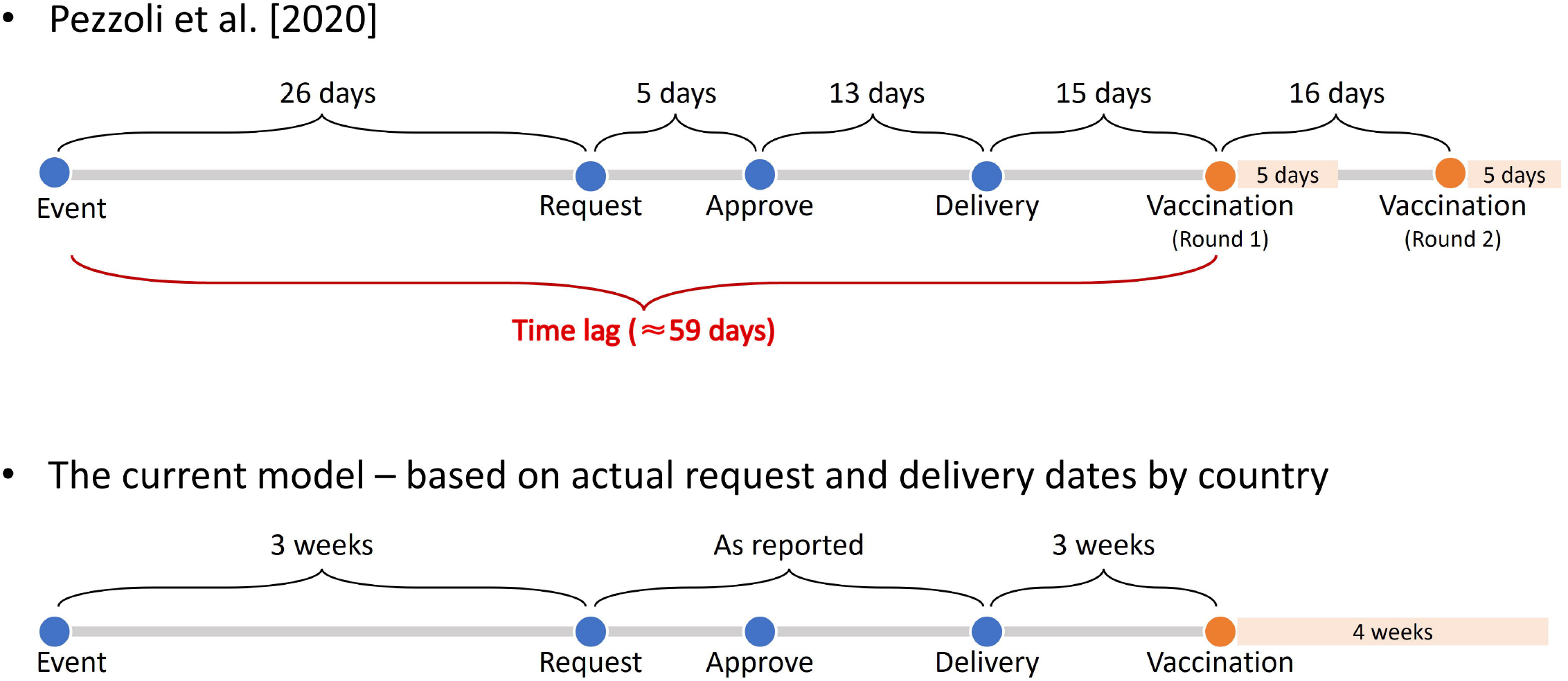
Time-lags until vaccination.

Vaccination impacts were estimated in terms of cases averted, deaths averted, and Disability-Adjusted Life Years (DALYs) averted compared to no vaccination and presented by scenario and country. The expected economic burden saved due to vaccination was also calculated by multiplying the number of cases averted by the estimated economic burden per cholera episode after inflating (or deflating) for the corresponding year (2023) using the GDP deflators^20, 21, 26^ (see Supplementary Table 5). The estimated economic burden was expressed in US dollars (US$) based on the alternative conversion factor which takes into account the official exchange rate diverging by an exceptionally large margin^26^, as well as in international dollars (I$) using the purchasing power parity (PPP).

### Sensitivity and uncertainty analysis

Considering a degree of uncertainty due to data-sparse settings, a series of sensitivity and uncertainty analyses were carried out. First, to address the uncertainty of the estimated parameters and model outcomes, the minimum and maximum values of reliable estimates ranging from 31 to 100 sets per country and meeting the sum of squared differences between the estimate and the observed incidence less than 0.001, were used to generate uncertainty intervals. Second, because there were multiple OCV requests per country over time, additional simulations were conducted assuming substantial OCV supplies at the time of the first outbreak and measuring the impact of one-time vaccination with varying coverage rates (50%, 70%, and 90%). These simulations were designed to evaluate if a one-time mass vaccination campaign with assumed coverage rates would have been effective in controlling outbreaks, compared to multiple OCV stockpile requests by country over time. If the actual coverage at the time of the first request exceeded the assumed coverage rate (50%, 70%, and 90%), the actual (higher) coverage was used. Third, given that the duration of an outbreak would affect the impact of vaccination and that time- and location-specific data for all countries were scarce, varying vaccination impacts were assessed based on shorter (8 weeks) and longer duration (19 weeks) scenarios of an outbreak, in addition to the default duration (12 weeks).^22^ Global percentage reduction of cases and 95% confidence intervals were generated by bootstrapping country-specific simulation outputs.

## RESULTS

The number of the OCV stockpile shipment is shown in Figure 2A. From 2013 to 2025, over 210 million doses and 61 million doses were shipped for reactive and preventive vaccinations, respectively. A total of 307 deliveries were recorded during the period, and 61 of the total deliveries were intended for preventive vaccinations. While there were continuous OCV shipments for reactive vaccinations during the entire study period, the OCV deliveries for preventive vaccinations were only reported from October 2016 to August 2022.

**Figure 2.**
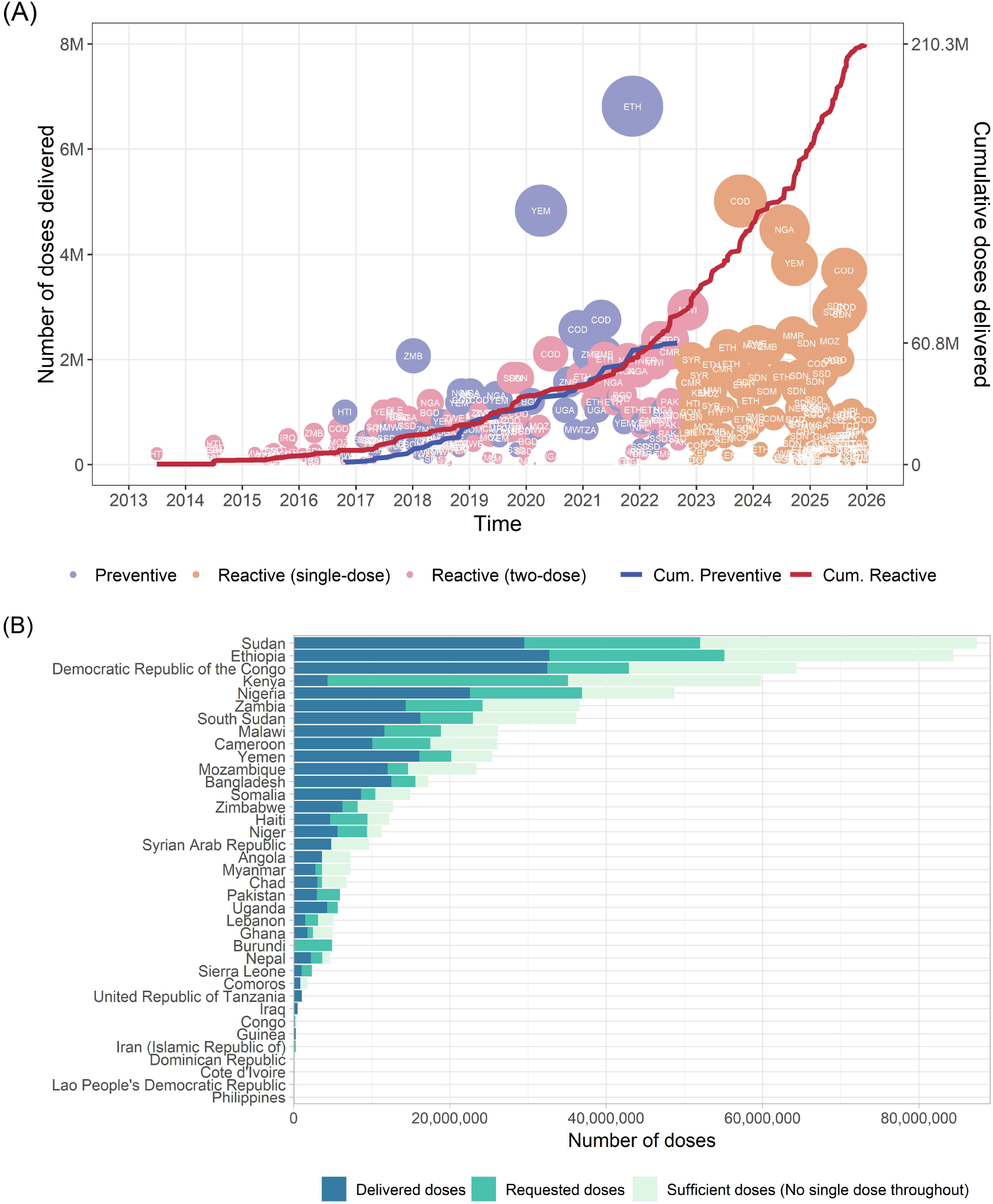
OCV stockpile shipment over time and the number of OCV doses by country. In (A), the size of each bubble corresponds to the size of doses delivered.

Figure 2B presents the number of OCV doses required by country: the actual number of doses delivered, the requested number of doses, and the sufficient number of doses meeting all country stockpile requests and sustaining the continuous two-dose vaccination scheme. A majority of the countries received a smaller number of OCVs than requested. In particular, countries including Kenya, Haiti, and Lebanon, etc. received less than 50% of doses than what they requested. Ethiopia received the highest number of OCVs. While DRC ranked the second in terms of the actual number of doses shipped, the requested number of doses was the second highest in Sudan following Ethiopia. A total number of OCVs increased from 552 million (requested doses) to 835 million (sufficient doses).

The three multipliers were estimated and applied to R_0_ to closely approximate the observed outbreaks in terms of the cumulative cases detected during the outbreak periods (see supplementary material for full details). Once the multipliers were determined, this procedure was repeated for all OCV recipient countries to approximate varying outbreak sizes based on every single request for reactive vaccination and its timing per country. The impact of vaccination is shown in Figure 3. With the actual OCV shipment scenario (scenario 1), vaccination prevented 8.1 million cases and 146,013 deaths which resulted in 6.73 million DALYs averted. If there were neither time lags nor the supply shortages meeting the stockpile requests and maintaining the continuous two-dose vaccination even after October 2022 (scenario 4), the vaccination impact would have been 17.4 million cases, 321,730 deaths, and 15.1 million DALYs averted. In terms of the number of cases averted, the vaccination impact was the greatest in Ethiopia under all scenarios as shown in Figure 3B. On the other hand, the number of averted deaths and DALYs was the largest in Sudan across all scenarios (see Supplementary Figures 13 and 14). This suggests that the magnitudes of vaccination impacts are not linearly proportionate to the absolute number of doses but variable by country depending on different levels of country-specific incidence, mortality, as well as coverage rates.

**Figure 3.**
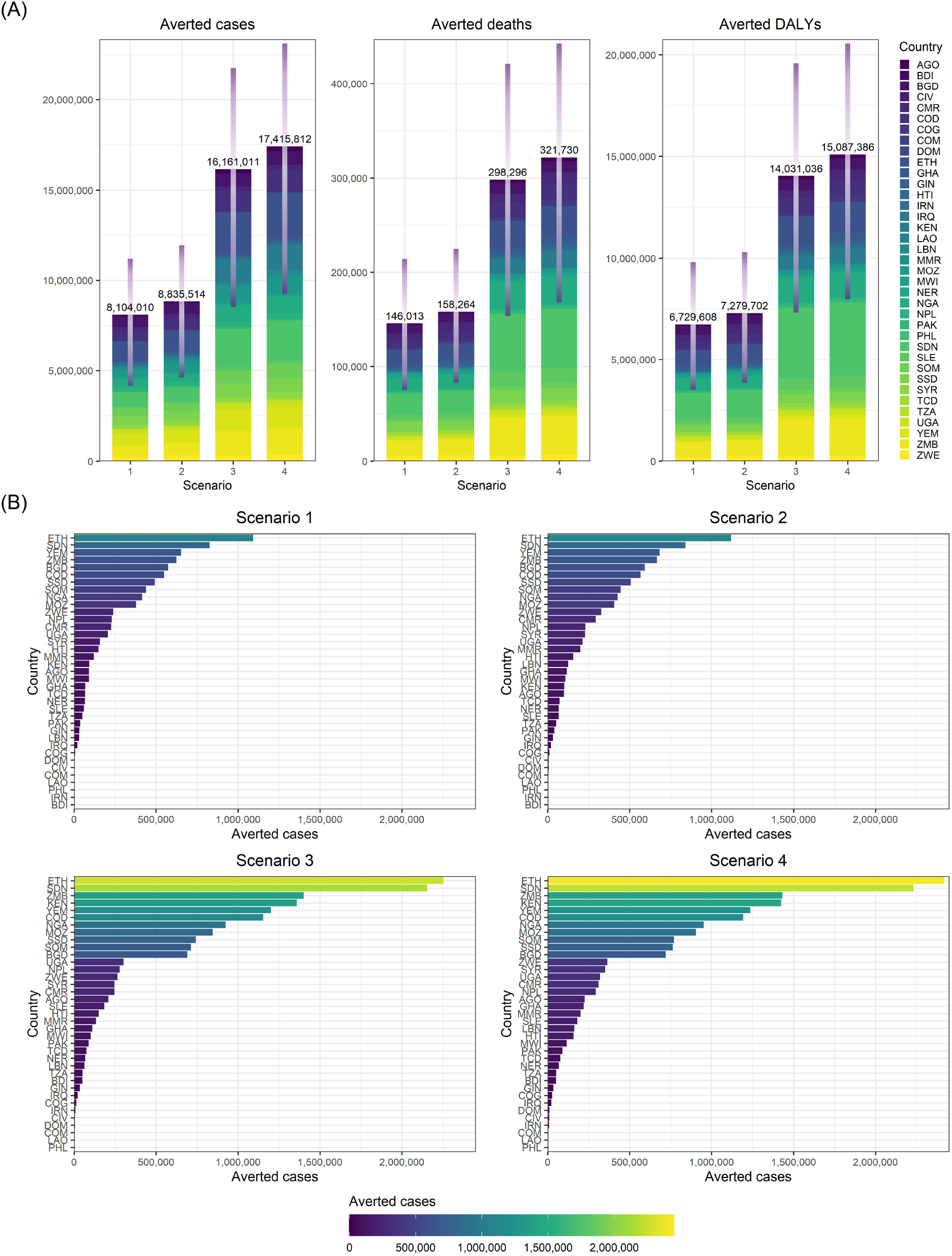
Impact of vaccination. (A) Aggregated impact of vaccination by scenario; (B) Number of cases averted by country

Figure 4A shows the percentage reduction due to vaccination by scenario. Overall, the percentage reduction increased from scenario 1 to scenario 4 as expected. From scenario 1 to 2, the additional benefit was achieved by changing the timing of vaccination and removing the time-lags for OCVs to arrive since the onset of an event. In other words, if the recipient countries had an in-country OCV stockpile and vaccinated during the early stage of an outbreak, a median of 22.8 % of case reduction would have been possible with no extra costs, compared to that of 15.5% reduction. It was also interesting to observe that while each of no time-lag scenario (scenario 2) and no supply shortage scenario (scenario 3) contributed to more reductions in cholera cases compared to scenario 1, vaccination impact was greater with the sufficient number of OCVs only (scenario 3) than with no time-lag alone (scenario 2), holding the other constraints constant.

**Figure 4.**
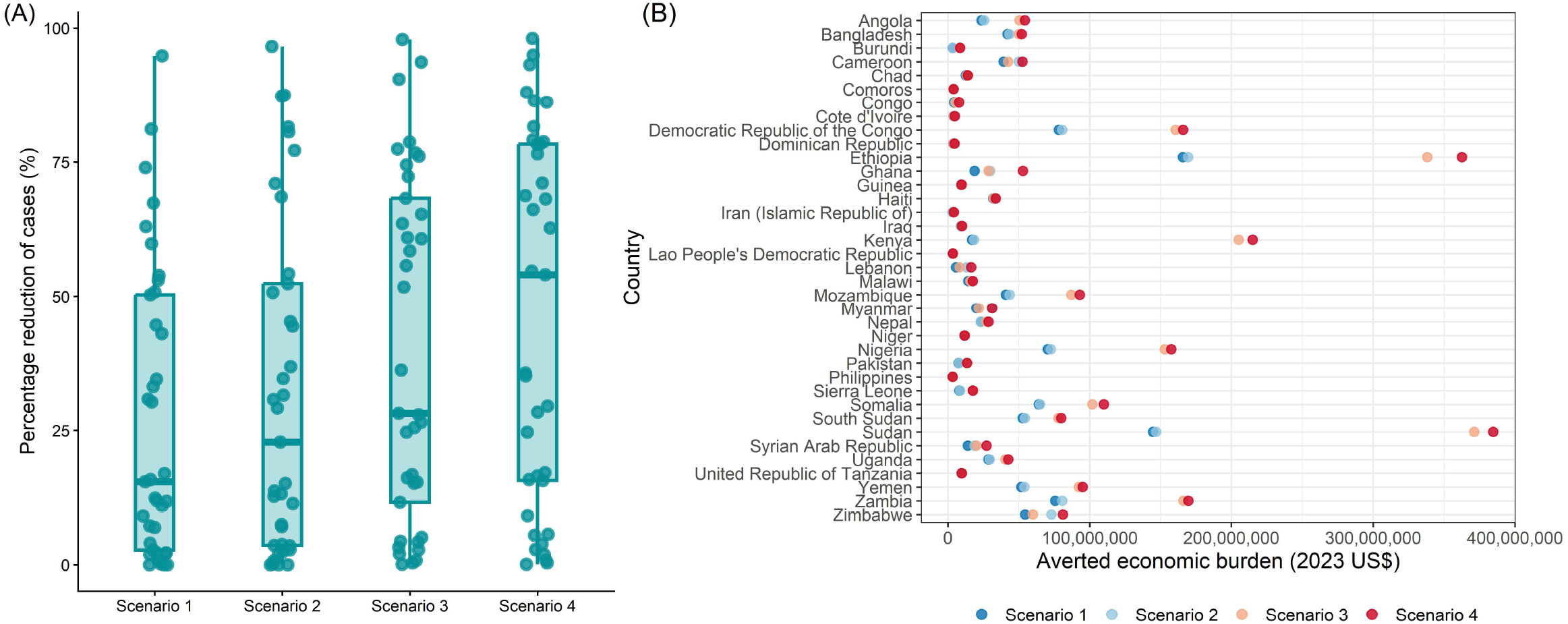
Percentage reduction due to vaccination and economic burden of cholera saved due to vaccination. See Supplementary Table 5 for country-specific economic burden values

The number of cases averted and percentage reduction due to vaccination were further demonstrated in Supplementary Figures 15 and 16 for alternative scenarios where a one-time mass vaccination campaign was assumed instead of multiple OCV requests per country. The percentage reduction was greater when vaccinating 50% of the risk population than scenario 4 and continued to increase as the coverage rate became higher. To address the uncertainty about time- and location-specific duration of an outbreak, additional simulation was carried out by varying the duration of an outbreak. As shown in Supplementary Figures 17, the resulting percentage reduction was the highest with an 8-week duration and the lowest with a 19-week duration across all four scenarios.

The economic burden of cholera saved due to the OCV stockpile is also shown in Figure 4B. The averted economic burden of cholera was the greatest in Sudan followed by Ethiopia under all four scenarios. This was mainly due to the larger number of averted cases and the higher economic burden of cholera per episode than other countries. The total averted economic burden of cholera was estimated to be around US$1.05 billion (I$3.21 billion), US$1.15 billion (I$3.54 billion), US$2.15 billion (I$6.64 billion), and US$2.33 billion (I$7.21 billion) under scenario 1, 2, 3, and 4, respectively.

## DISCUSSION

The current study quantified the contributions of the OCV stockpile to the global health through the development of the low-cost vaccine and tech-transfers since its inception in 2013. Our model estimated that from 2013 to the end of 2025, both reactive and preventive vaccination campaigns implemented through the global OCV stockpile averted more than 6.7 million DALYs and 146,000 deaths in 37 recipient countries. In addition, the vaccination saved US$1.05 billion of the economic burden due to cholera.Despite this significant contribution of the OCV stockpile to limit the impact of cholera, the delay in delivery of vaccine to affected communities and the supply shortage of OCVs due to the unpredictable nature of the emergency-based demand made it challenging to optimize the potential benefits. Because of the time lags, the recipient countries often missed the optimal timing of vaccination during an outbreak situation. Moreover, the supply shortage led to a reduced and sub-optimal scale of response as compared to the countries’ stockpile requests. The requirement of the single-dose vaccination regime since October 2022 further blunted the potential vaccine impact. The current model demonstrates the potential impact of the OCV stockpile without delays in campaign implementation and/or supply shortages to guide future decision-making processes.

The additional benefits estimated by shifting scenarios from 1 to 2 highlight the importance of the timing of vaccination and have implications not only for the current OCVs but also for future cholera vaccines.For example, for countries with a robust cold-chain system in place, the OCV stockpile can be stored within a country in advance and used to increase the effectiveness of vaccination through a well-planned preventive campaign or by intervening during the early stage of an outbreak reducing the spread of the disease (transmission) in a more efficient manner. Importantly, compared to the existing practice, such benefits do not necessarily come with additional costs associated with vaccine supplies which are known to be the major cost driver for a vaccination campaign.^27, 28^

The greater benefit observed under scenario 3 provides insights into the future policy-making process. Currently, all stockpile recipient countries are required to implement a single-dose OCV campaign due to the shortage of OCVs. According to our findings, the continuous support for a two-dose vaccination campaign would bring much larger benefits than the single-dose vaccination scheme. This is mainly because cholera is highly prevalent among young children under 5 years, and a single-dose vaccination with the OCV is known to be much less efficacious among young children than a two-dose vaccination.^6,7^

Some areas of uncertainty deserve attention. First, due to the scarcity of the observed longitudinal data, it was challenging to estimate the accurate size of various cholera outbreaks that each country had experienced over time since 2013. The current study estimated the multipliers based on varying outbreak sizes from a set of the observed data and applied to other countries with no data to approximate outbreak sizes. While each outbreak size and its corresponding multiplier were determined to closely reflect the observed number of cases, for countries with no observed data, the requested number of OCVs was used as a proxy to determine relative outbreak sizes corresponding to the multipliers (see full details in supplementary material). The estimation would be more robust if there were empirically observed longitudinal data in more countries. Second, the proportion of populations at risk was estimated by utilizing the composite index developed for TRF. While the risk factors used for the TRF index are applicable for cholera as well, future research is required to update the risk factors related to water and sanitation. Third, climate indicators (temperature and precipitation) were also incorporated into the model, but understanding the association with cholera occurrences was limited due to the absence of robust longitudinal data and specific geographical locations within a country. Fourth, a time lag from the onset of each outbreak to a country stockpile request to the ICG would likely vary but is difficult to measure for every single outbreak for all recipient countries in an accurate manner. While the current model assumed the average time lag to be 3 weeks similar to a previous study^16^, the vaccination impact would likely change with varying time lags as shown in the current study as well (scenario 2).Nonetheless, it is worth noting that the current study adopted the actual (observed) time lags from the date of request to the date of delivery to increase the accuracy of the estimation. Lastly, it is worth noting that the uncertainties created by the lack of primary data would likely cause biases in different directions. For example, the intensity of an outbreak would likely be reduced after vaccination but might still be sustained longer than expected, which would result in the overestimation of the current vaccination impact. The sensitivity analysis showed that vaccination impact was the greatest when the duration of an outbreak was assumed to be short as expected. In addition, given the absence of a granular level of empirical data in low- and middle-income countries settings, the vaccination impact would be underestimated (or overestimated) depending upon the difference between the high-risk populations estimated in the study and the actual number of populations affected by outbreaks. Thus,the current study further estimated the changes in vaccination impact by varying vaccination coverage rates.

While OCV has been an effective intervention tool to control cholera in many parts of the world, there are shortcomings as well. For example, the current OCV has a relatively short shelf-life, requires cold-chain equipment, and provides suboptimal protection for children younger than 5 years. It should be noted that other cholera vaccine candidates are currently in development. The capsule-type thermostable cholera vaccine called Duochol is in its early stage but expected to have a longer shelf-life with no cold-chain requirement and to confer a higher efficacy including among children younger than 5 years compared to the current OCV. Thus, if successful, the vaccine will likely enable in-country stockpile capacity resulting in more rapid response as well as making it easier to reach remote areas (hard-to- reach populations), which in turn, might lead to more effective cholera controls. Moreover, a Cholera Conjugate Vaccine successfully completed its phase 1 trial in 2024. Injectable conjugate vaccines elicit long lasting T-cell dependent immune responses in young children which may provide a longer duration of protection, providing an opportunity for vaccination to be incorporated into EPI reducing the burden of repeated vaccination campaigns.

## CONCLUSION

The current study found that the impact of the WHO stockpile on global health was significant. This successful initiative was possible due to continuous collaborative efforts by a number of international partners including the development of the low-cost OCV and tech-transfers. It should be noted that a greater supply of OCV and its more extensive use in national cholera control campaigns will further reduce outbreaks and meaningfully contribute to the WHO’s plan to reduce cholera deaths by 90% by 2030. Additional OCV products are expected to be available in the near future which will expand the supply and reduce the possibility of shortages. Future research is needed to understand the optimal usage and potential impacts of multiple cholera vaccines which can be administered by different strategies (i.e., routine or campaign) at varying time points, geographical locations, and among varied age cohorts to maximize the impact and cost-effectiveness (value for money) of public interventions.

## Data Availability

All data are available from the GTFCC OCV database: https://apps.epicentremsf.org/public/app/gtfcc.

## Acknowledgements

This study was initiated and carried out by authors’ in-kind contributions (the Policy and Economic Research (PER) department at the International Vaccine Institute (IVI)). The authors appreciate and acknowledge the partial funding support from the IVI Europe Regional Office through investment from the Swedish Ministry for Foreign Affairs grant UD2022/000123.

